# Immunosuppressive regimens and long-term kidney transplant outcomes: a dual survival modeling framework

**DOI:** 10.64898/2025.12.03.25341568

**Authors:** Kunle Timothy Apanisile, Meng-Hao Li, Hadi El-Amine, Naoru Koizumi

**Author notes:** Corresponding author (KTA).

## Abstract

Optimizing immunosuppressive therapy remains central to improving long-term outcomes after kidney transplantation. Both induction and maintenance therapies are widely used, yet their comparative effectiveness across diverse recipient populations requires further evaluation. This national retrospective cohort study analyzed 228,855 deceased-donor kidney transplant recipients using data from 2000-2024. Multivariable Cox proportional hazards (PH) models were used for clinical inference, and four machine learning (ML) survival models: random survival forest (RSF), support vector machine (SVM), penalized Cox regression (CoxNet), and extreme gradient boosting optimized with the Cox partial likelihood (XGBoost-Cox), were developed to assess predictive performance for death-censored graft failure and all-cause patient mortality. Model performance was evaluated using the concordance index (C-index) and time-dependent area under the curve (tdAUC).

Maintenance regimens incorporating calcineurin inhibitors (CNI) and mycophenolate mofetil (MMF) were associated with lower hazards for both graft failure (CNI+MMF: hazard ratio [HR] 0.72, 95% confidence interval [CI] 0.70-0.74; CNI+MMF+steroids: HR 0.84, 95% CI 0.82-0.87) and patient mortality (CNI+MMF: HR 0.78, 95% CI 0.76-0.81; CNI+MMF+steroids: HR 0.90, 95% CI 0.88-0.93). Among induction therapies, antithymocyte globulin (ATG) was associated with lower hazards for both outcomes, whereas interleukin-2 receptor (IL-2R) antagonists and Alemtuzumab demonstrated neutral associations. Combined ATG+IL-2R therapy was associated with higher hazard of graft failure (HR 1.09). Recipient diabetes, dialysis dependence, older age, and higher Kidney Donor Profile Index were strong adverse predictors. Traditional Cox regression achieved robust discrimination (graft failure C-index: 0.685; patient mortality C-index: 0.704), comparable to ML survival models.

These findings support the continued association of CNI and MMF maintenance regimens with favorable long-term transplant outcomes while demonstrating variation across induction strategies. The dual analytical framework integrating classical Cox modeling with ML survival methods, suggests that Cox models remain highly competitive for clinical inference, whereas ML approaches provide complementary predictive value to support individualized post-transplant risk stratification.

## Introduction

Kidney transplantation is the gold standard treatment for kidney failure, offering superior survival, cost-effectiveness, and quality of life compared to long-term dialysis [1,2]. The long-term success of a transplant, however, is threatened by the recipient’s immune response to the foreign graft. This response is primarily triggered by the disparity in human leukocyte antigens (HLAs) between the donor and recipient, which activates the recipient’s T cells and can lead to graft failure through two principal pathways: antibody-mediated and cellular-mediated rejection [3]. Antibody-mediated rejection involves B-cells producing donor-specific antibodies, a process dependent on CD4+ T-cell help [4,5]. In contrast, cellular rejection is driven directly by the cytotoxic activity of T cells, alongside other immune cells such as macrophages and natural killer cells [6]. Without tailored immunosuppressive medications to modulate the immune response, a persistent immune attack leads to progressive immune-mediated injury, marked by inflammation, tissue damage, and potential graft failure [7]. Such injury may present as acute rejection early after transplantation or evolve into chronic rejection over time, thereby highlighting the vital role of immunosuppressive treatment.

Immunosuppressive therapies aim to prevent or control this immune-mediated injury by suppressing the recipient’s immune system. By doing so, they reduce the immune response against the allograft, attenuate inflammation, and help preserve its function and viability. Therefore, post-transplant immunosuppression is essential for limiting immune-mediated injury and maintaining long-term graft function.

Despite the crucial role of immunosuppressive therapy, it carries potential risks and side effects, emphasizing the need to balance immune suppression and complications. The definitive protocol remains undetermined [8-10], with most centers employing an approach involving induction therapy with interleukin-2 receptor (IL-2R) antibodies or antithymocyte globulin (ATG), along with a maintenance regimen comprising steroids, calcineurin inhibitors (CNIs), and mycophenolate mofetil (MMF) [11,12]. Although these advances have substantially improved short-term graft survival, the challenge of achieving durable long-term outcomes persists [13-15]. Therefore, ongoing research continues to seek regimens that maximize short-term benefits while minimizing the risk of long-term deterioration [16].

Predicting long-term outcomes is fundamental to personalizing care and improving graft survival. For decades, survival analysis methods, particularly Cox proportional hazards (PH) regression, have been the cornerstone of outcome evaluation in kidney transplantation. Cox models allow estimation of hazard ratios (HRs), providing clinicians with interpretable measures of the relative risk of graft failure or patient death associated with specific therapies or clinical covariates [17-19]. These models are well-suited to accommodate censoring, varying follow-up times, and the multifactorial nature of post-transplant outcomes.

At the same time, the growing availability of large-scale transplant datasets has created opportunities for machine learning (ML) methods to complement traditional survival models. More recently, ML survival models such as random survival forests (RSFs), support vector machines (SVMs), and gradient boosting survival models, have demonstrated the potential to improve risk prediction in transplantation by modeling nonlinear interactions and high-dimensional patterns that may not be fully represented in standard Cox regression [20-22]. Yet, many prior ML studies in kidney transplantation have been limited by small sample sizes, older cohorts, or lack of direct comparison with classical survival models using the same covariate structure [23-26]. Additionally, few have explicitly examined both induction and maintenance immunosuppressive strategies within ML frameworks, and even fewer have evaluated their performance against death-censored graft survival and patient mortality outcomes using contemporary national data.

The current study addresses these gaps through analysis of a large and recent national cohort of deceased-donor kidney transplant recipients and examining both induction and maintenance immunosuppressive therapies in relation to long-term outcomes. Death-censored graft survival and all-cause patient mortality were evaluated using multivariable Cox proportional hazards models to provide clinically interpretable estimates of relative risk associated with specific therapeutic regimens. In parallel, multiple ML survival models were developed using the same covariate structure to determine whether they improve predictive performance beyond classical Cox regression. Model performance was evaluated using concordance index (C-index) and time-dependent area under the curve (tdAUC) metrics at clinically relevant follow-up intervals, enabling direct comparison of inference-focused and prediction-focused approaches within a unified analytical framework.

By integrating classical survival modeling with contemporary ML survival methods in a large and up-to-date national dataset, this study provides a rigorous evaluation of immunosuppressive regimen effectiveness while simultaneously assessing the extent to which advanced predictive algorithms offer added clinical value. This dual analytical perspective is intended to support clinically informed regimen selection and the advancement of individualized post-transplant risk stratification.

## Materials and methods

### Data source and study population

This retrospective cohort study used data from the United Network for Organ Sharing (UNOS) registry. Two data extracts were provided: an earlier file covering January 1, 2000, through May 29, 2021, and a subsequent update containing transplants from January 1, 2015, through October 30, 2024. These datasets were merged using transplant recipient identifiers and procedure-level record keys, with harmonization of variable definitions and removal of duplicate entries during overlapping years, to create a unified dataset spanning January 1, 2000, to October 30, 2024. The analysis was restricted to adult recipients of deceased-donor kidney transplants, identified using the donor type variable. Living-donor kidney transplants were excluded, as were observations with incomplete or non-positive follow-up times as well as extensive missing values for some critical variables. The final analytic cohort included 228,855 deceased-donor kidney transplant recipients.

#### Study outcome

Two clinically relevant post-transplant outcomes were examined. The primary endpoint was death-censored graft failure, defined as the time from kidney transplantation to return to dialysis or re-transplantation, with deaths occurring in the presence of a functioning graft treated as censored events at the time of death. The secondary endpoint was all-cause patient mortality, defined as time from transplantation to death from any cause, regardless of graft function. Follow-up time for both outcomes was calculated from transplantation date to event occurrence or administrative censoring on October 30, 2024, whichever occurred first. Both outcomes were analyzed using time-to-event methods appropriate for right-censored data.

#### Predictor variables and data preprocessing

Candidate predictors encompassed donor, recipient, and transplant characteristics, including demographic, clinical, and immunologic factors (e.g., age, Kidney Donor Profile Index [KDPI], HLA mismatches, calculated panel-reactive antibody [cPRA]). Immunosuppressive regimens were the key focus. ATG constituted the predominant induction therapy in the cohort (59.6%, n = 136,458), with IL-2R antagonists (19.4%, n = 44,512) and Alemtuzumab (12.2%, n = 27,982) used less frequently. For maintenance therapy, a triple regimen of CNI, MMF, and steroids predominated (67.1%, n = 153,515), while a steroid-free combination of CNI + MMF accounted for 24.3% (n = 55,637). Other regimens were used less frequently.

Missing data were handled with a combination of complete-case analysis and imputation. The proportion of missing data was low (<1%) for most variables. Continuous variables (e.g., serum creatinine, body mass index [BMI]) were imputed with medians, and categorical variables (e.g., diabetes) were imputed with modes. Missing cPRA values (14.8%) were imputed as 0 and accompanied by a binary missingness indicator. This approach preserves cohort size while allowing the model to adjust for potential non-random missingness. The empirical distribution of cPRA in the study cohort demonstrated a median value of 0%, supporting this imputation strategy and reflecting registry-wide patterns. This approach is further supported by the historical registry structure, where cPRA reporting was incomplete or inconsistently captured in earlier transplant eras due to evolving Organ Procurement and Transplantation Network (OPTN) data collection practices. Cold ischemic time (1.4%) was imputed with the median and similarly flagged.

### Statistical analysis

#### Descriptive and survival analysis

Continuous variables were presented as medians with interquartile ranges (IQRs), whereas categorical measures were reported as counts and corresponding percentages. Kaplan–Meier survival curves were generated for graft and patient survival, including subgroup comparisons by donor and recipient factors (e.g., donation after circulatory death [DCD] vs. non-DCD, expanded criteria donor [ECD], KDPI quartiles, recipient diabetes). Group differences in survival were assessed using the log-rank test.

#### Multicollinearity assessment and model specification

Prior to multivariable modeling, variance inflation factors (VIF) were calculated to assess multicollinearity among candidate predictors. A systematic approach was employed where categorical variables with inherent collinearity (geographic sharing, recipient race, donor race) were coded using reference category exclusion. Geographic sharing utilized ’local’ as reference, while race variables used ’white’ as reference category for both recipient and donor. Variables with VIF exceeding 10 were iteratively removed until all remaining predictors demonstrated acceptable collinearity levels (VIF < 10). Following the addition of transplant era indicators to the final models, VIFs were re-assessed; all covariates, including the era indicators, remained below the multicollinearity threshold (VIF < 10). Continuous variables were maintained in their natural units to preserve clinical interpretability of HRs.

#### Cox regression

A multivariable Cox PH model was fitted separately for death-censored graft failure and all-cause patient mortality. All models were constructed using the final predictor set identified after assessing and addressing multicollinearity among covariates. To account for secular changes in transplant practice, immunosuppressive protocols, and allocation policy over the study period (2000–2024), models were additionally adjusted for transplant era (2000–2004, 2005–2009, 2010–2014, 2015–2019, and 2020–2024), with the earliest era serving as the reference category. The final era category was defined to capture contemporary transplant practice, including recent allocation system updates and current immunosuppressive management patterns. Because treatment allocation was not randomized, models were designed to estimate adjusted associations rather than causal treatment effects. HRs with 95% confidence intervals (CIs) were estimated. The PH assumptions were assessed using Schoenfeld residuals and graphical diagnostics. Sensitivity analyses refitting models without transplant era adjustment yielded qualitatively similar effect estimates, supporting the robustness of the primary findings (S1 and S2 Tables). Final models balanced clinical interpretability and statistical parsimony.

### Machine learning survival models

Multiple ML survival algorithms were implemented using identical predictor variables to enable direct comparison with classical Cox regression. The ensemble included: RSF with memory-optimized implementation using warm-start and batch training to prevent computational overload; SVM with kernel optimization; extreme gradient boosting optimized with the Cox partial likelihood (XGBoost-Cox); and penalized Cox regression (CoxNet) with elastic net regularization. Data were randomly split into training (75%) and testing (25%) sets stratified by event status to maintain outcome distribution. Model parameters were selected based on established defaults and transplantation literature, with computational efficiency considerations for the large-scale national dataset. Continuous predictors were standardized for algorithms requiring feature scaling (CoxNet, SVM), while RSF and XGBoost-Cox utilized natural units.

#### Model evaluation

Each model’s predictive accuracy was assessed using the independent test set not used during training. The concordance index (C-index) was used to evaluate overall discrimination. Time-dependent Area Under the Curve (tdAUC) was computed at 1-, 3-, 5-, and 10-years post-transplant, and the mean tdAUC was used as a summary measure of longitudinal predictive performance. All analyses were performed in Python 3.13.7 using lifelines, scikit-survival, and scikit-learn libraries. This study was reported in accordance with the STROBE reporting guideline for observational cohort studies (S4 Checklist).

## Results

### Cohort characteristics

The study cohort comprised 228,855 recipients of deceased-donor kidney transplants from 2000-2024. Recipients had a median age of 55 years (interquartile range [IQR] 44-63), with 60.4% male and 36.3% with diabetes. Racial distribution included 39.5% White, 33.5% Black, 17.8% Hispanic, and 7.0% Asian recipients. Diabetes was present in 36.3% of recipients, and 83.0% were receiving dialysis at the time of transplantation. The median BMI was 27.8 kg/m² (IQR 24.2–32.0), and the median cPRA was 0% (IQR 0–29). Approximately 12.2% were retransplants, and the median waiting time prior to transplantation was 639 days (IQR 193–1314).

Donor characteristics reflected a median age of 41 years (IQR 29–52), with 61.3% male donors. DCD accounted for 22.2% of transplants, while 16.2% met expanded criteria donor (ECD) definitions. The median KDPI was 40% (IQR 20–62), and the median donor creatinine was 1.0 mg/dL (IQR 0.7–1.4). DCD donors had higher KDPI compared with non-DCD donors (median [IQR] 50% [33–68] vs 37% [17–60]; Mann–Whitney p<0.001), reflecting expected differences in donor selection and organ utilization patterns (S3 Table).

Immunosuppressive regimens were heterogeneous: ATG was the most common induction agent (59.6%), followed by IL-2R (19.4%) and Alemtuzumab (12.2%). For maintenance therapy, the majority received a triple regimen of CNI + MMF + steroids (67.1%), with an additional 24.3% maintained on CNI + MMF without steroids. Full characteristics are presented in ***Table 1***.

**Table 1.** Baseline characteristics of the transplant cohort.

| Characteristic | Value |
| --- | --- |
| Study cohort | 228,855 recipients |
| **Recipient characteristics** |  |
| Age (years), median (IQR) | 55.0 (44.0–63.0) |
| Male | 138,269 (60.4%) |
| White | 90,383 (39.5%) |
| Black | 76,765 (33.5%) |
| Hispanic | 40,658 (17.8%) |
| Asian | 16,069 (7.0%) |
| Retransplant | 27,819 (12.2%) |
| On dialysis | 189,966 (83.0%) |
| Diabetes | 82,998 (36.3%) |
| BMI (kg/m<sup>2</sup>), median (IQR) | 27.8 (24.2–32.0) |
| cPRA (%), median (IQR) | 0.0 (0.0–29.0) |
| Wait time (days), median (IQR) | 639.0 (193.0–1314.0) |
| Creatinine (mg/dL), median (IQR) | 8.0 (5.9–10.5) |
| **Donor characteristics** |  |
| Age (years), median (IQR) | 41.0 (29.0–52.0) |
| Male donor | 140,221 (61.3%) |
| White donor | 157,516 (68.8%) |
| Black donor | 29,915 (13.1%) |
| Hispanic donor | 32,546 (14.2%) |
| Asian donor | 5,525 (2.4%) |
| DCD (%) | 50,907 (22.2%) |
| ECD (%) | 37,092 (16.2%) |
| Donor diabetes | 18,102 (7.9%) |
| Donor hypertension | 66,879 (29.2%) |
| BMI (kg/m<sup>2</sup>), median (IQR) | 27.1 (23.5–31.7) |
| KDPI, median (IQR) | 40% (20–62%) |
| Donor creatinine (mg/dL), median (IQR) | 1.0 (0.7–1.4) |
| **Induction regimen** |  |
| ATG | 136,458 (59.6%) |
| IL-2R | 44,512 (19.4%) |
| Alemtuzumab | 27,982 (12.2%) |
| ATG + IL-2R | 8,226 (3.6%) |
| No induction | 11,677 (5.1%) |
| **Maintenance regimen** |  |
| CNI + MMF + Steroids | 153,515 (67.1%) |
| CNI + MMF | 55,637 (24.3%) |
| Other/atypical regimens | 19,703 (8.6%) |
| Cold ischemia time (hours), median (IQR) | 18.0 (12.9–23.1) |
| **Transplant factors** |  |
| Local allocation (%) | 147,075 (64.3%) |
| Regional allocation (%) | 34,192 (14.9%) |
| National allocation (%) | 47,588 (20.8%) |

Values are presented as median interquartile range (IQR) or n (%), as appropriate. Abbreviations: ATG = anti-thymocyte globulin; IL-2R = interleukin-2 receptor; CNI = calcineurin inhibitor; MMF = mycophenolate mofetil; KDPI = Kidney Donor Profile Index; DCD = donation after circulatory death; ECD = expanded criteria donor; BMI = body mass index; cPRA = calculated panel-reactive antibody.

### Overall graft and patient survival

During a median follow-up of 4.0 years (IQR 1.9–7.0), there were 55,346 death-censored graft failures (24.2%) and 43,772 all-cause patient deaths (19.1%) among 228,855 deceased-donor kidney transplant recipients. The distribution of events, censoring, and follow-up time is summarized in ***Table 2***.

**Table 2.** Event and Follow-Up Summary.

| Outcome | N (analysis set) | Events n (%) | Censored n (%) | Median follow-up, years (IQR) |
| --- | --- | --- | --- | --- |
| **Graft failure (death-censored)** | 228,855 | 55,346 (24.2) | 173,509 (75.8) | 4.0 (1.9–7.0) |
| **Patient mortality (all-cause)** | 228,855 | 43,772 (19.1) | 185,083 (80.9) | 4.3 (2.0–7.2) |

Events and censoring represent follow-up status and should not be interpreted as Kaplan–Meier survival probabilities. Follow-up duration was calculated from the date of transplantation to event occurrence or administrative censoring.

Kaplan–Meier analyses demonstrated progressive decline in both graft and patient survival over time, with relatively high early post-transplant survival that gradually decreased during long-term follow-up (***Table 3, Figs 1–2***). The estimated death-censored graft survival was 95.7% at 1 year, 88.5% at 3 years, 80.1% at 5 years, and 57.8% at 10 years, corresponding to a median graft survival of approximately 12.0 years (95% CI, 11.9–12.0). In comparison, patient survival was 96.8% at 1 year, 91.4% at 3 years, 85.1% at 5 years, and 66.5% at 10 years, with a median all-cause survival of approximately 15.0 years.

**Fig 1.**
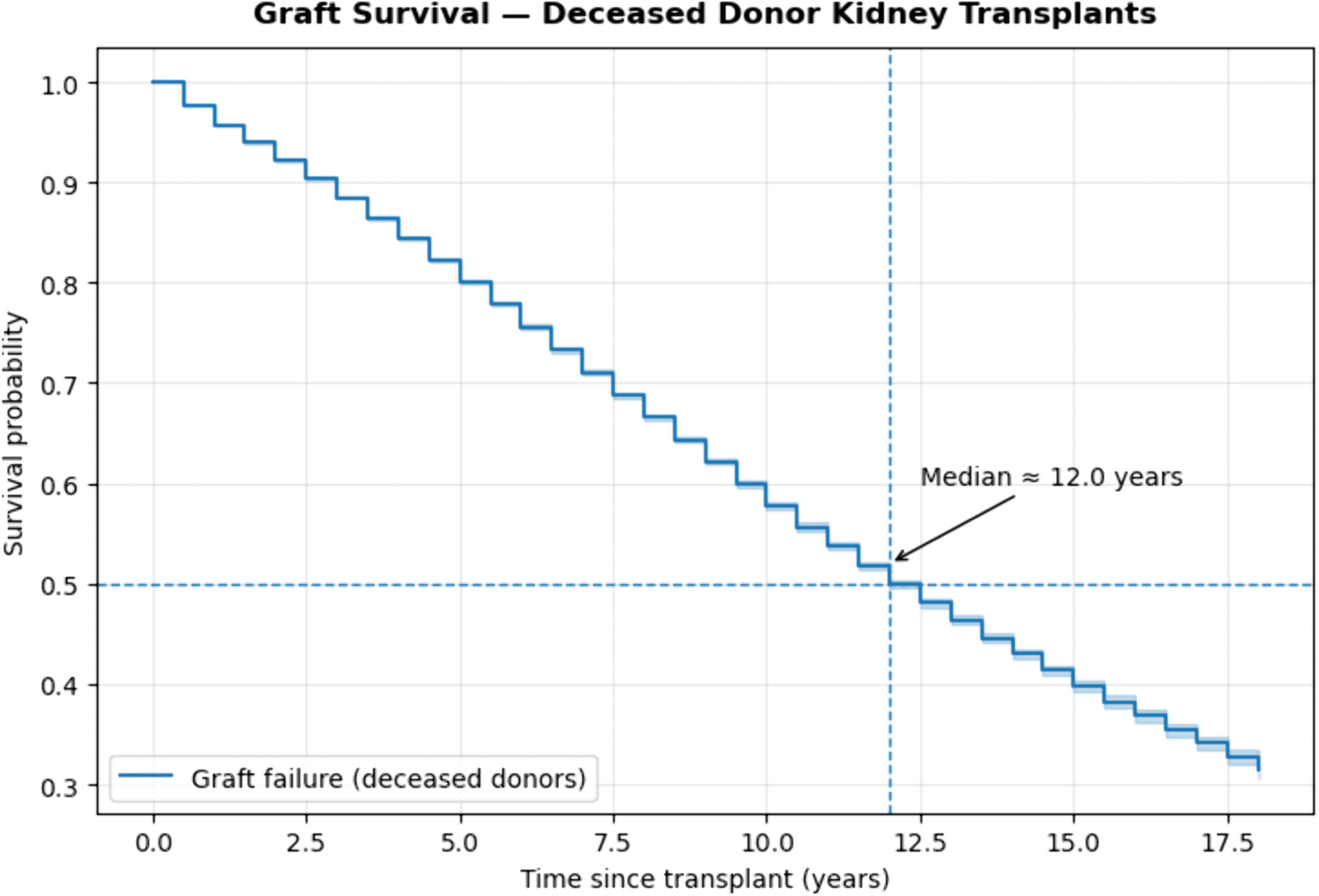
Kaplan–Meier curve for death-censored graft survival among deceased-donor kidney transplants.

**Fig 2.**
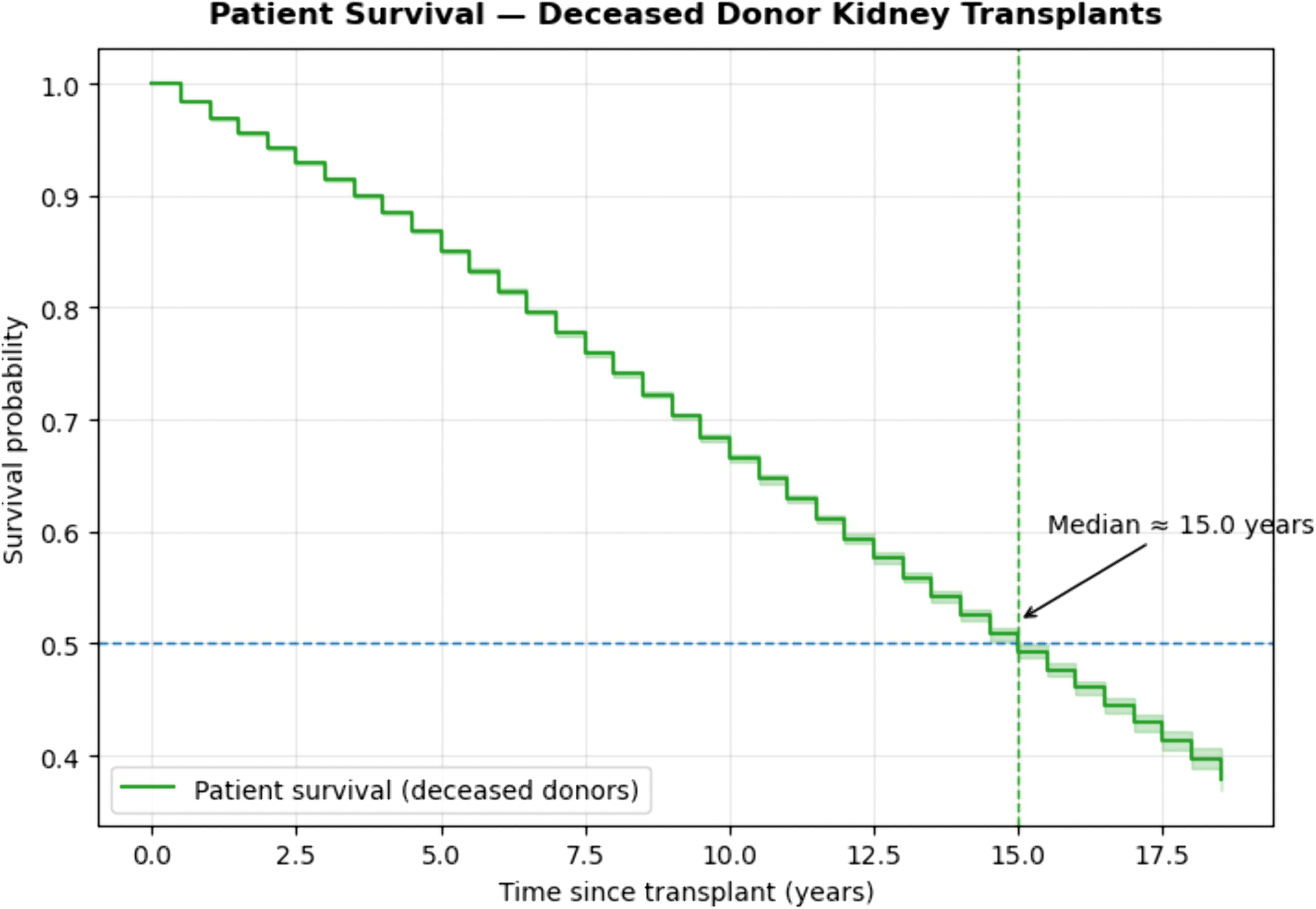
Kaplan–Meier curve for all-cause patient survival among deceased-donor kidney transplants.

**Table 3.** Kaplan–Meier Survival Estimates at Selected Time Points.

| **Time (years)** | **N at risk – Graft** | **Graft survival (95% CI)** | **N at risk – Patient** | **Patient survival (95% CI)** |
| --- | --- | --- | --- | --- |
| 1 | 197,478 | 0.957 (0.956–0.958) | 201,096 | 0.968 (0.968–0.969) |
| 3 | 140,017 | 0.885 (0.883–0.886) | 146,458 | 0.914 (0.913–0.916) |
| 5 | 94,936 | 0.801 (0.799–0.803) | 101,904 | 0.851 (0.849–0.852) |
| 10 | 20,935 | 0.578 (0.574–0.581) | 23,944 | 0.665 (0.662–0.669) |

Values represent Kaplan–Meier survival probabilities derived from time-to-event analysis at specific post-transplant time points with 95% confidence intervals. CI = confidence interval. Median graft survival ≈ 12.0 years; median patient survival ≈ 15.0 years. N at risk indicates the number of recipients remaining under observation and event-free at each interval.

### Subgroup survival analyses

Graft and patient survival differed significantly across key donor and recipient subgroups (***Figs 3–10***). Recipients of kidneys from DCD showed a statistically significant but clinically small difference in graft survival compared with non-DCD transplants (median 12.1 vs 12.0 years; log-rank χ² = 4.87, p = 0.027). Differences in patient survival were more pronounced, with DCD recipients exhibiting a shorter median survival (14.3 vs 14.9 years; log-rank χ² = 40.11, p < 0.001). In contrast, kidneys from ECD showed markedly reduced outcomes. Median graft survival was 8.1 years for ECD versus 13.0 years for non-ECD donors (log-rank χ² = 4150.03, p < 0.001), and median patient survival was 10.4 vs 15.8 years, respectively (log-rank χ² = 2961.72, p < 0.001).

**Fig 3.**
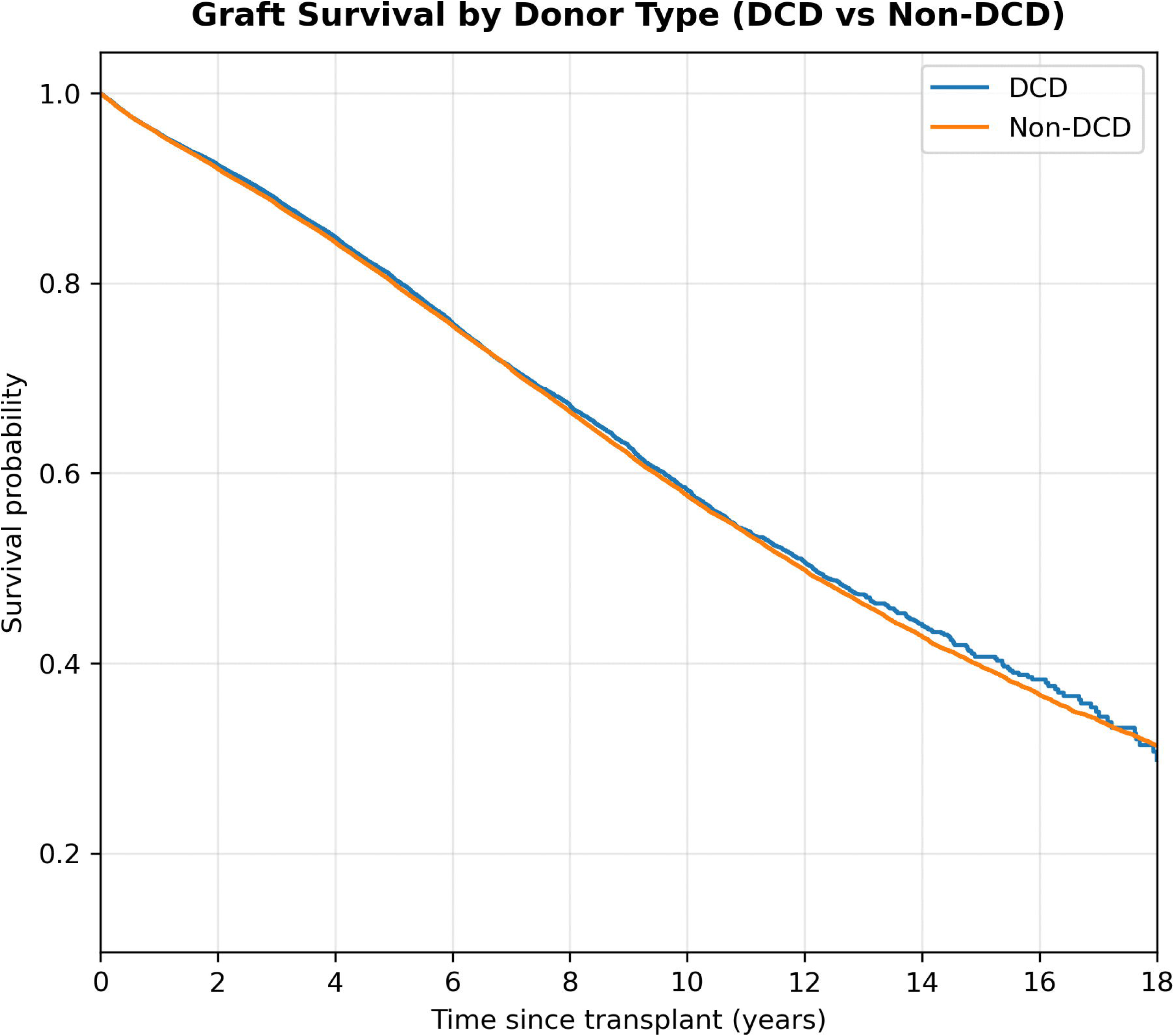
Graft survival by donor type (DCD vs non-DCD). Kaplan–Meier curves comparing graft survival between DCD and non-DCD kidney transplants. Log-rank χ² and *p*-values were derived from univariate comparisons.

**Fig 4.**
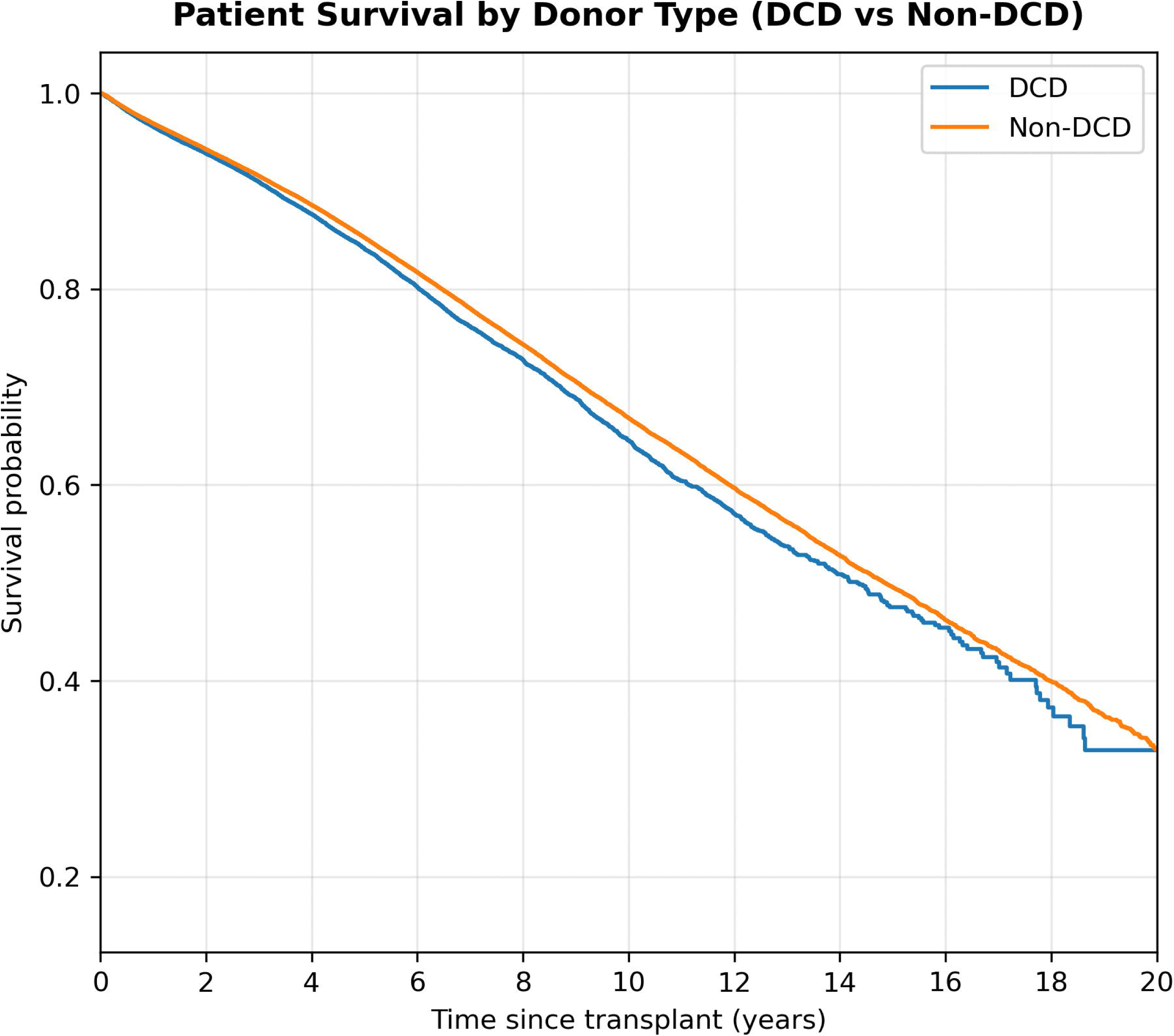
Patient survival by donor type (DCD vs non-DCD). Kaplan–Meier curves comparing patient survival between DCD and non-DCD kidney transplants. Log-rank χ² and *p*-values were derived from univariate comparisons.

**Fig 5.**
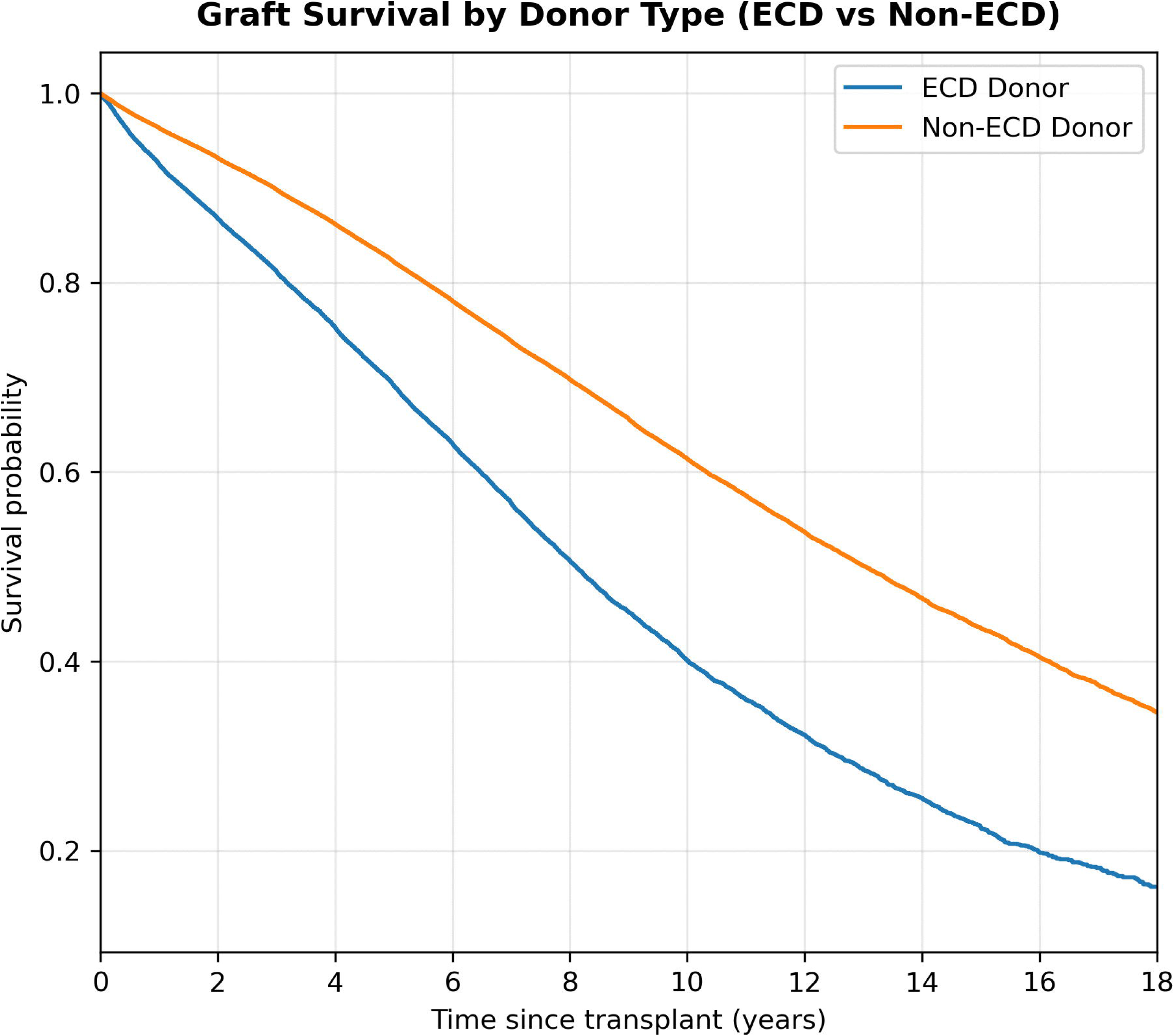
Graft survival by donor quality (ECD vs non-ECD). Kaplan–Meier curves comparing graft survival between expanded criteria donors (ECD) versus non-ECD kidney transplants. Log-rank χ² and *p*-values were derived from univariate comparisons.

**Fig 6.**
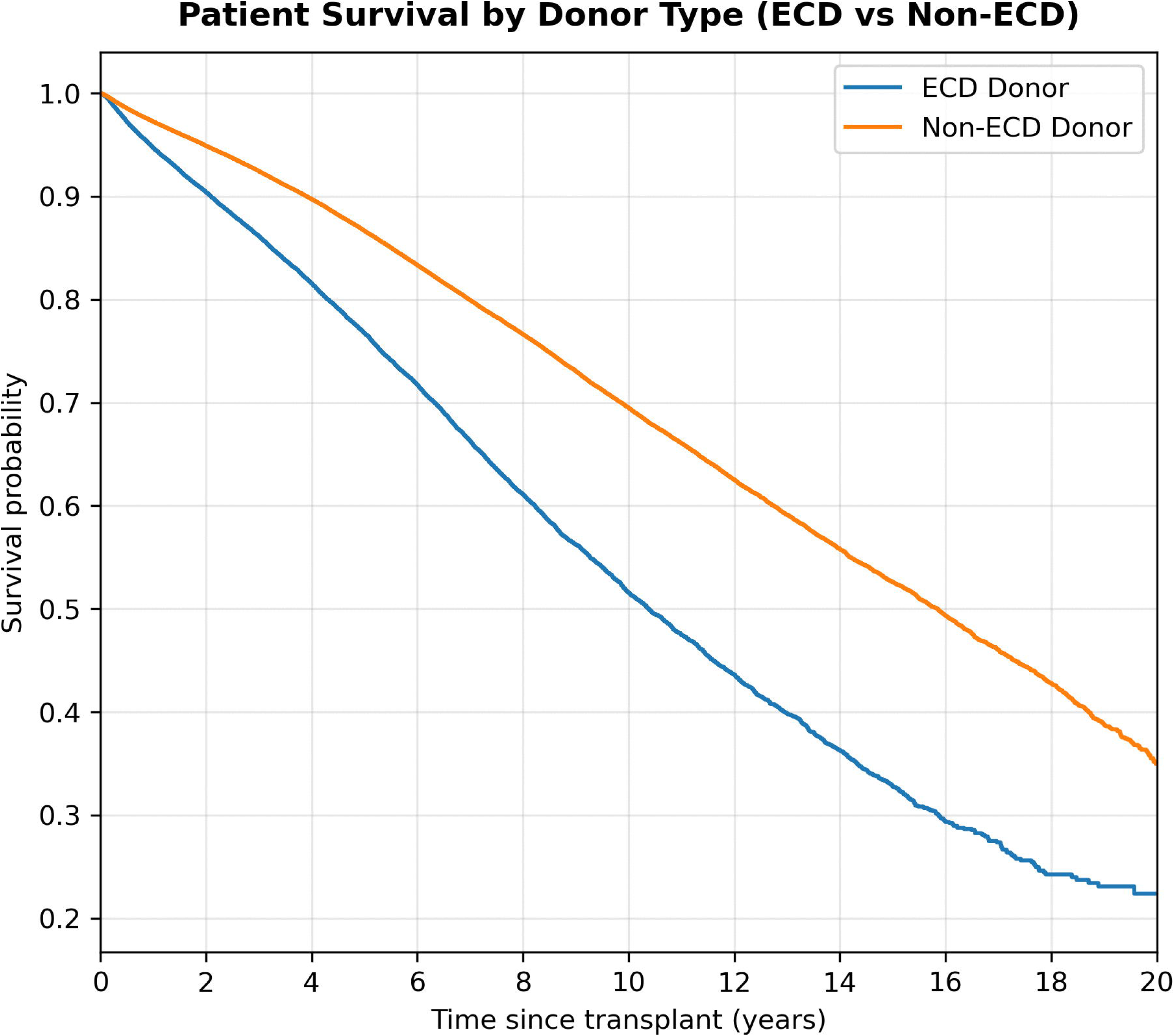
Patient survival by donor quality (ECD vs non-ECD). Kaplan–Meier curves comparing patient survival between ECD versus non-ECD kidney transplants. Log-rank χ² and *p*-values were derived from univariate comparisons.

**Fig 7.**
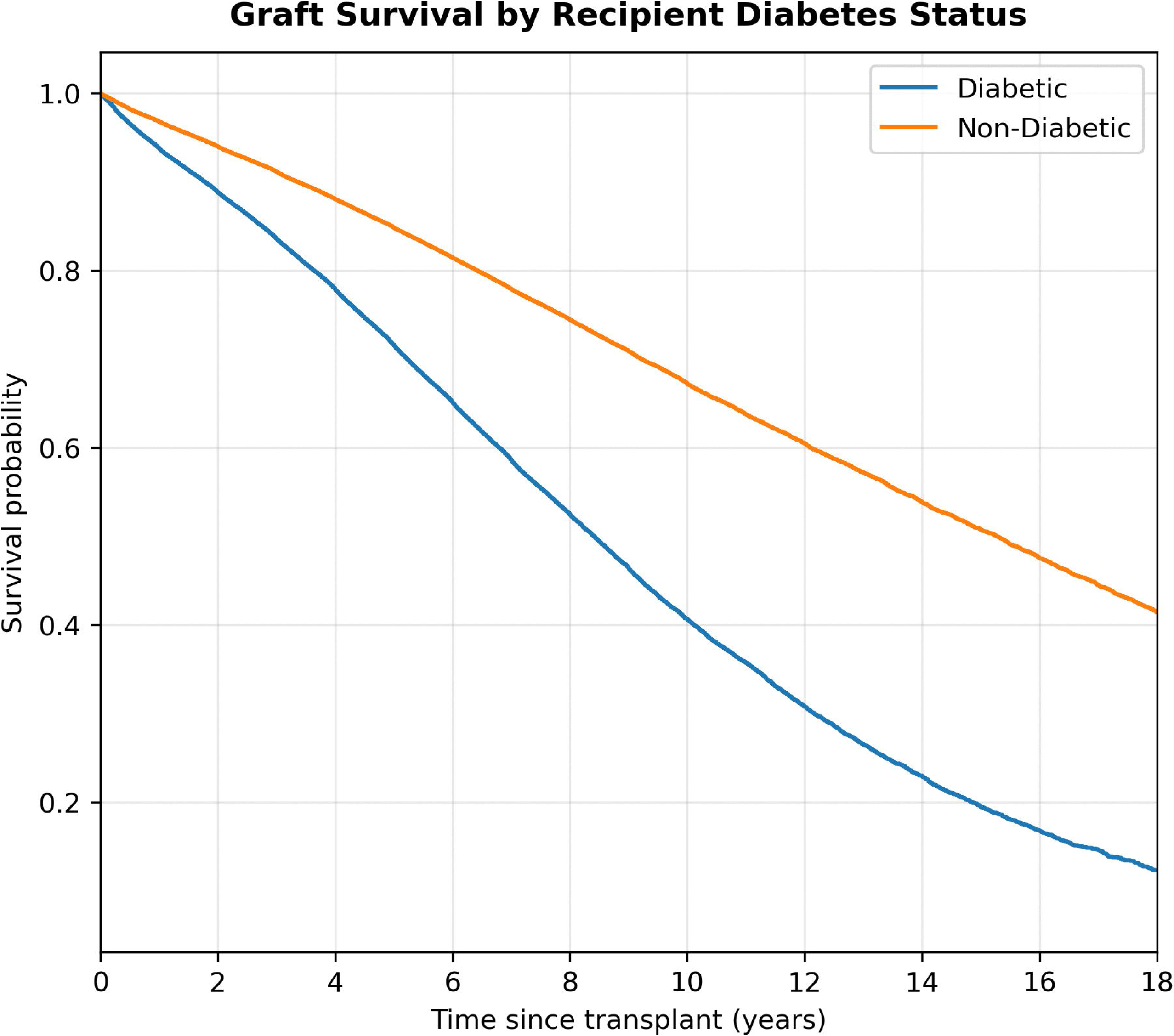
Graft survival by recipient diabetes status. Kaplan–Meier curves comparing graft survival between diabetic and non-diabetic recipients at the time of transplantation. Log-rank χ² and *p*-values were derived from univariate comparisons.

**Fig 8.**
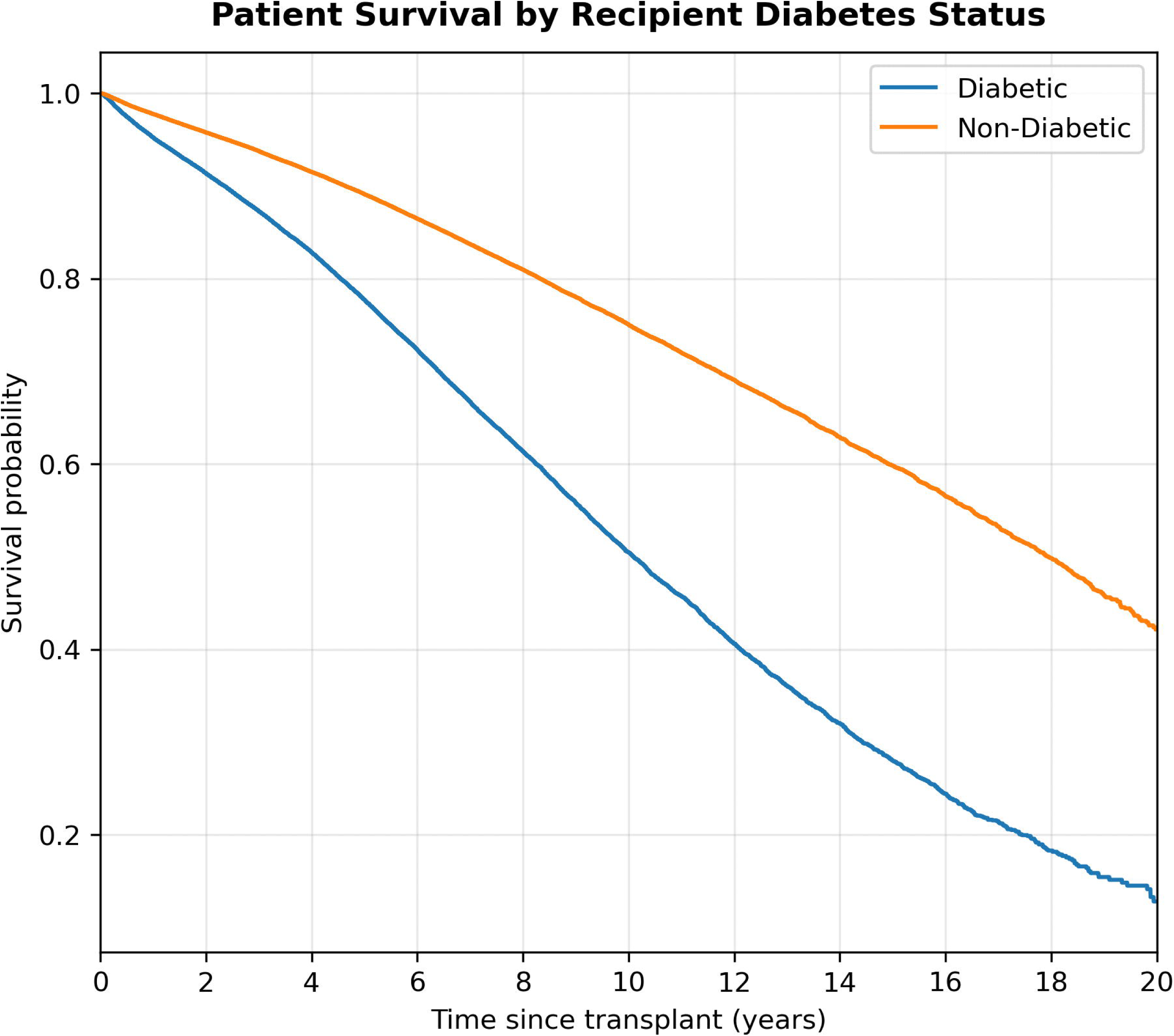
Patient survival by recipient diabetes status. Kaplan–Meier curves comparing patient survival between diabetic and non-diabetic recipients. Log-rank χ² and *p*-values were derived from univariate comparisons.

**Fig 9.**
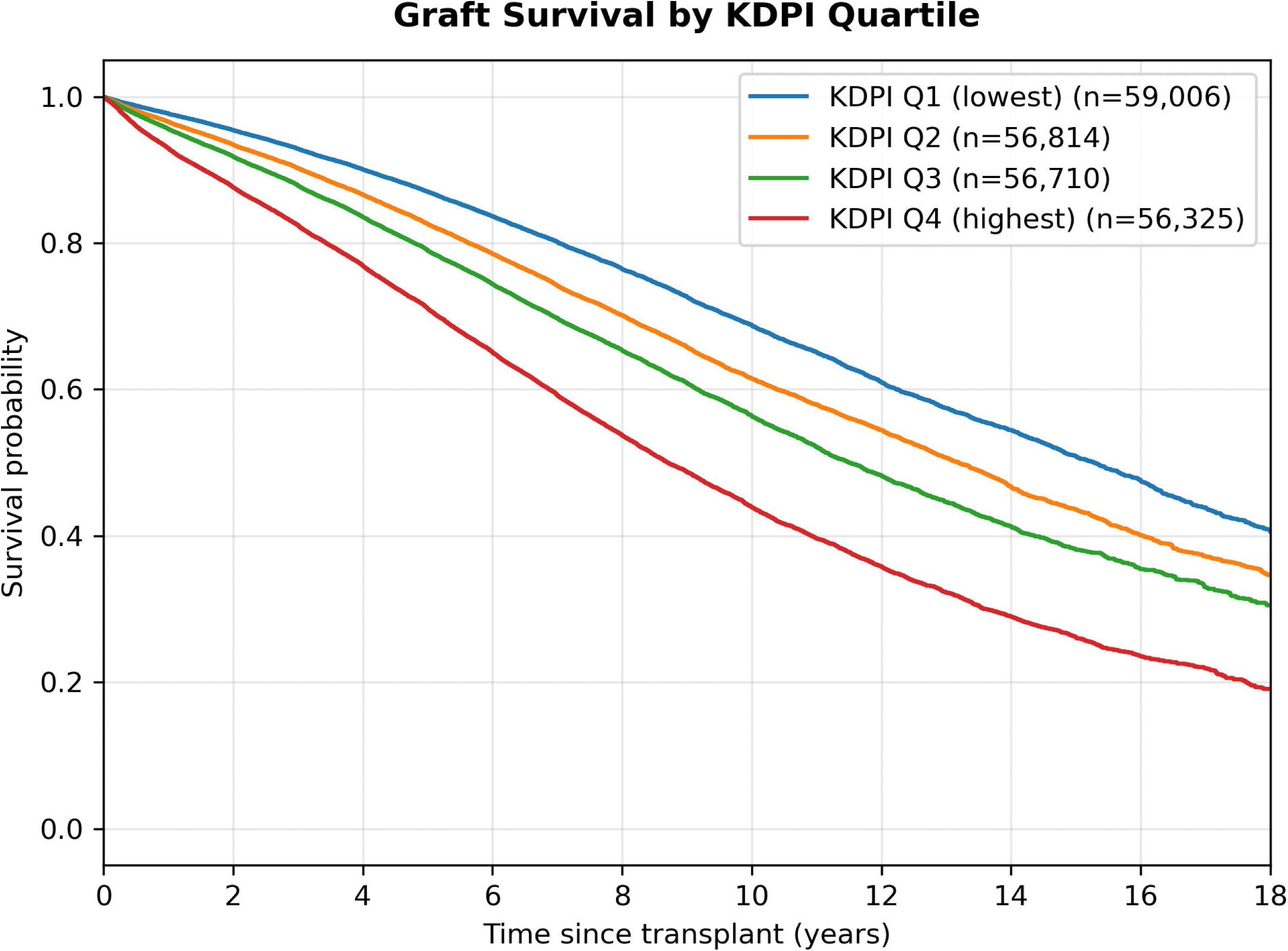
Graft survival by KDPI quartiles. Kaplan–Meier curves stratified by KDPI quartiles (Q1–Q4). Log-rank χ² and *p*-values were derived from a multigroup comparison.

**Fig 10.**
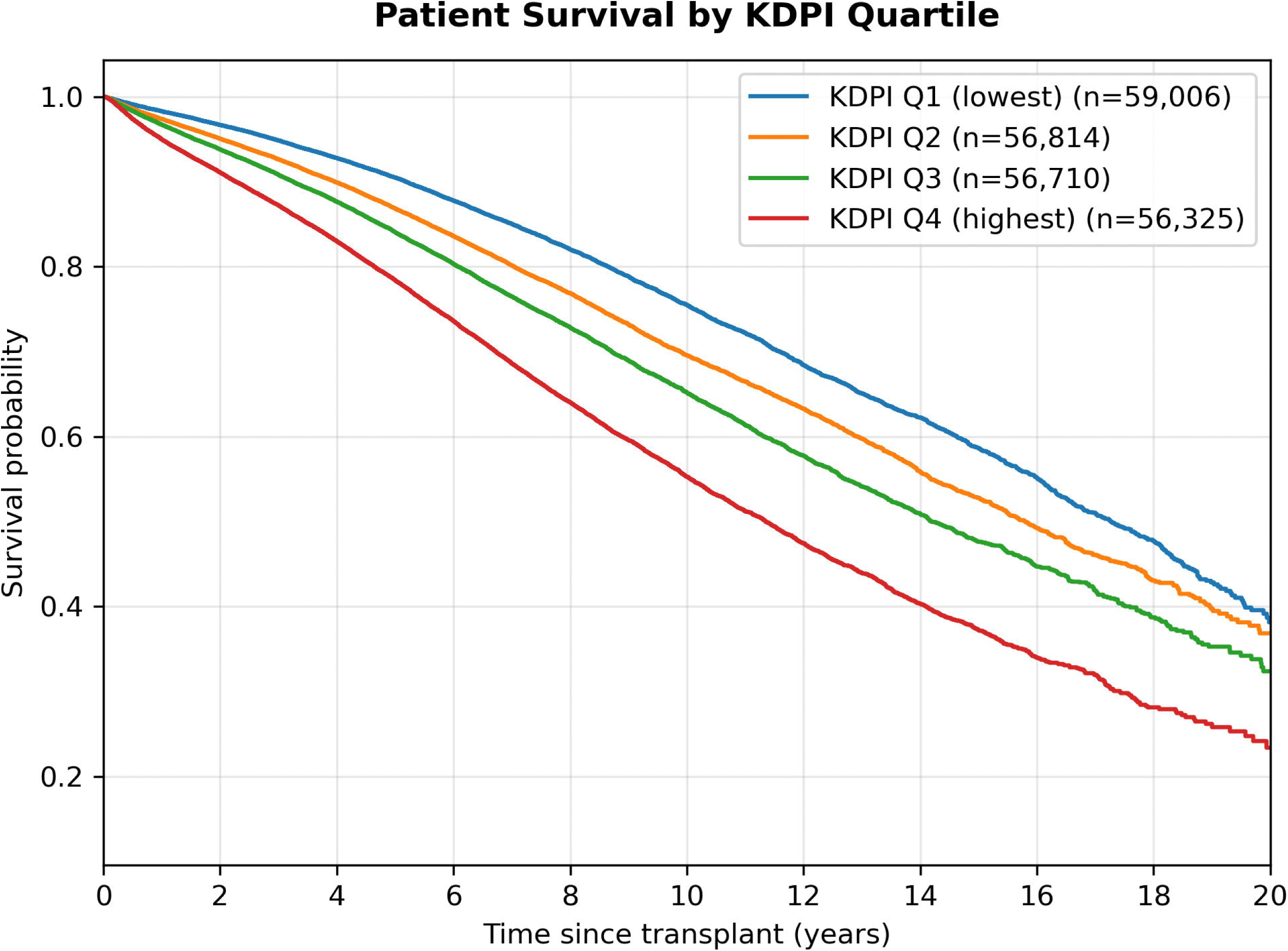
Patient survival by KDPI quartiles. Kaplan–Meier curves stratified by KDPI quartiles (Q1–Q4). Log-rank χ² and *p*-values were derived from a multigroup comparison.

Recipient diabetes was associated with striking survival disparities. Graft survival differed sharply by diabetic status, with recipients who had diabetes experiencing a median survival of 8.4 years, whereas those without diabetes reached 15.3 years (log-rank χ² = 8463.18, p < 0.001). Median patient survival followed a similar pattern (10.1 vs 17.9 years; log-rank χ² = 7871.70, p < 0.001). The KDPI quartiles showed a stepwise gradient (***Figs 9–10***). Graft and patient survival declined progressively from the lowest KDPI quartile (Q1) to the highest (Q4) (multigroup log-rank χ² = 5420.45 and 3746.92, respectively; p < 0.001 for both).

### Multivariable Cox proportional hazards models

Multivariable Cox regression models identified distinct risk factor profiles for graft failure and patient mortality while controlling for comprehensive donor, recipient, and transplant characteristics (***Table 4-5***). Immunosuppressive regimens demonstrated significant associations with both outcomes, though with varying effect magnitudes across therapeutic classes. For maintenance immunosuppression, CNI + MMF regimens were associated with substantially lower hazards of both graft failure (HR 0.72, 95% CI 0.70-0.74, p < 0.001) and patient mortality (HR 0.78, 95% CI 0.76-0.81, p < 0.001). The addition of steroids to this backbone in triple-therapy regimens (CNI + MMF + steroids) was also associated with lower hazards for both outcomes, although with more modest effect sizes (graft failure HR 0.84, 95% CI 0.82-0.87; patient mortality HR 0.90, 95% CI 0.88-0.93; both p < 0.001).

**Table 4.** Multivariable Cox proportional hazards model for graft failure.

| Covariate | HR (95% CI) | P-value |
| --- | --- | --- |
| **Immunosuppression** |  |  |
| CNI + MMF (no steroids) | 0.72 (0.70–0.74) | <0.001 |
| CNI + MMF + steroids | 0.84 (0.82–0.87) | <0.001 |
| ATG induction | 0.93 (0.89–0.97) | 0.002 |
| IL-2R induction | 1.04 (0.99–1.09) | 0.101 |
| IL-2R + ATG induction | 1.09 (1.03–1.16) | 0.002 |
| Alemtuzumab induction | 0.97 (0.92–1.02) | 0.234 |
| **Recipient factors** |  |  |
| Age (per year) | 1.04 (1.04–1.04) | <0.001 |
| Male | 1.15 (1.13–1.18) | <0.001 |
| Black race | 0.97 (0.95–0.99) | 0.010 |
| White race | Reference | – |
| Hispanic ethnicity | 0.74 (0.72–0.76) | <0.001 |
| Asian race | 0.61 (0.59–0.64) | <0.001 |
| Diabetes | 1.63 (1.60–1.66) | <0.001 |
| Retransplant | 1.27 (1.24–1.31) | <0.001 |
| On dialysis at transplant | 1.57 (1.53–1.61) | <0.001 |
| Serum creatinine at transplant (mg/dL) | 0.97 (0.97–0.97) | <0.001 |
| BMI (per kg/m<sup>2</sup>) | 1.00 (1.00–1.01) | <0.001 |
| cPRA (per %) | 1.00 (1.00–1.00) | <0.001 |
| Wait time (per day) | 1.00 (1.00–1.00) | <0.001 |
| **Donor factors** |  |  |
| Age (per year) | 1.00 (1.00–1.00) | <0.001 |
| Male | 1.00 (0.98–1.01) | 0.633 |
| White race | Reference | – |
| Black race | 1.08 (1.05–1.11) | <0.001 |
| Hispanic ethnicity | 0.96 (0.94–0.99) | 0.006 |
| Asian race | 1.01 (0.95–1.06) | 0.829 |
| Diabetes | 1.05 (1.02–1.09) | 0.002 |
| Hypertension | 1.00 (0.98–1.03) | 0.726 |
| BMI (per kg/m<sup>2</sup>) | 1.00 (1.00–1.00) | 0.010 |
| Serum creatinine (mg/dL) | 0.99 (0.98–1.00) | 0.027 |
| ECD donor | 1.01 (0.98–1.04) | 0.478 |
| DCD donor | 0.91 (0.89–0.93) | <0.001 |
| KDPI (per 10 units) | 1.68 (1.57–1.81) | <0.001 |
| **Transplant factors** |  |  |
| Cold ischemia time (per hour) | 1.00 (1.00–1.00) | <0.001 |
| Local allocation | Reference | – |
| Regional allocation | 1.00 (0.97–1.03) | 0.968 |
| National allocation | 1.00 (0.98–1.03) | 0.713 |

Abbreviations: ATG = anti-thymocyte globulin; IL-2R = interleukin-2 receptor; CNI = calcineurin inhibitor; MMF = mycophenolate mofetil; ECD = expanded criteria donor; DCD = donation after circulatory death; KDPI = Kidney Donor Profile Index; BMI = body mass index; cPRA = calculated panel reactive antibody. Hazard ratios adjusted for donor, recipient, and transplant factors.

**Table 5.** Multivariable Cox proportional hazards model for patient mortality.

| Covariate | HR (95% CI) | P-value |
| --- | --- | --- |
| **Immunosuppression** |  |  |
| CNI + MMF (no steroids) | 0.78 (0.76–0.81) | <0.001 |
| CNI + MMF + steroids | 0.90 (0.88–0.93) | <0.001 |
| ATG induction | 0.93 (0.89–0.98) | 0.004 |
| IL-2R induction | 0.95 (0.91–1.00) | 0.070 |
| IL-2R + ATG | 1.02 (0.96–1.09) | 0.470 |
| Alemtuzumab induction | 0.95 (0.90–1.01) | 0.103 |
| **Recipient factors** |  |  |
| Age (per year) | 1.05 (1.05–1.05) | <0.001 |
| Male | 1.19 (1.17–1.22) | <0.001 |
| Black race | 0.92 (0.90–0.94) | <0.001 |
| White race | Reference | – |
| Hispanic ethnicity | 0.76 (0.74–0.78) | <0.001 |
| Asian race | 0.64 (0.61–0.66) | <0.001 |
| Diabetes | 1.67 (1.63–1.70) | <0.001 |
| Retransplant | 1.22 (1.18–1.26) | <0.001 |
| On dialysis at transplant | 1.54 (1.50–1.58) | <0.001 |
| Serum creatinine at transplant (mg/dL) | 0.97 (0.96–0.97) | <0.001 |
| BMI (per kg/m<sup>2</sup>) | 1.00 (1.00–1.01) | <0.001 |
| cPRA (per %) | 1.00 (1.00–1.00) | <0.001 |
| Wait time (per day) | 1.00 (1.00–1.00) | <0.001 |
| **Donor factors** |  |  |
| Age (per year) | 1.00 (1.00–1.00) | <0.001 |
| Male | 0.99 (0.97–1.01) | 0.563 |
| White race | Reference | – |
| Black race | 1.05 (1.02–1.09) | 0.006 |
| Hispanic ethnicity | 0.98 (0.95–1.01) | 0.111 |
| Asian race | 1.01 (0.95–1.08) | 0.722 |
| Diabetes | 1.05 (1.01–1.09) | 0.013 |
| Hypertension | 1.04 (1.01–1.06) | 0.004 |
| BMI (per kg/m<sup>2</sup>) | 1.00 (1.00–1.00) | 0.421 |
| Serum creatinine (mg/dL) | 1.00 (0.99–1.01) | 0.420 |
| ECD donor | 0.99 (0.96–1.03) | 0.740 |
| DCD donor | 1.01 (0.98–1.04) | 0.488 |
| KDPI (per 10 units) | 1.37 (1.26–1.48) | <0.001 |
| **Transplant factors** |  |  |
| Cold ischemia time (per hour) | 1.00 (1.00–1.00) | <0.001 |
| Local allocation | Reference | – |
| Regional allocation | 1.03 (1.00–1.06) | 0.037 |
| National allocation | 1.02 (0.99–1.04) | 0.261 |

Abbreviations: ATG = anti-thymocyte globulin; IL-2R = interleukin-2 receptor; CNI = calcineurin inhibitor; MMF = mycophenolate mofetil; ECD = expanded criteria donor; DCD = donation after circulatory death; KDPI = Kidney Donor Profile Index; BMI = body mass index; cPRA = calculated panel reactive antibody. Hazard ratios adjusted for donor, recipient, and transplant factors.

Among induction therapies, ATG was associated with lower hazards for both graft failure (HR 0.93, 95% CI 0.89-0.97, p = 0.002) and patient mortality (HR 0.93, 95% CI 0.89-0.98, p = 0.004). IL-2R showed neutral association with graft failure (HR 1.04, 95% CI 0.99-1.09, p = 0.101) and patient mortality (HR 0.95, 95% CI 0.91-1.00, p = 0.070). Alemtuzumab demonstrated neutral effects for both outcomes (graft failure HR 0.97, 95% CI 0.92-1.02, p = 0.234; patient mortality HR 0.95, 95% CI 0.90-1.01, p = 0.103). Combination therapy with ATG + IL-2R was associated with increased hazard for graft failure (HR 1.09, 95% CI 1.03-1.16, p = 0.002) but showed neutral association with patient mortality (HR 1.02, 95% CI 0.96-1.09, p = 0.470).

Recipient diabetes was among the strongest predictors of adverse outcomes, increasing the risk of graft failure by 63 % (HR 1.63, 95 % CI 1.60–1.66, p < 0.001) and patient mortality by 67 % (HR 1.67, 95 % CI 1.63–1.70, p < 0.001). Older recipient age was also independently associated with higher hazards for both graft failure (HR 1.04 per year, 95 % CI 1.04–1.04) and mortality (HR 1.05 per year, 95 % CI 1.05–1.05; both p < 0.001). Being on dialysis at the time of transplantation conferred approximately 1.5-fold greater risk for both outcomes. Both retransplantation (graft HR 1.27; mortality HR 1.22) and male recipient (graft HR 1.15; mortality HR 1.19) showed increased risks for each outcome. By contrast, recipients identified as Asian or Hispanic ethnicity in registry data experienced significantly lower hazards for both graft loss and death compared with White recipients (graft: HR 0.61 and 0.74; patient: HR 0.64 and 0.76, all p < 0.001). Black recipients had a modestly lower adjusted hazard of graft failure (HR 0.97, 95% CI 0.95-0.99, p = 0.010) and a more pronounced lower adjusted hazard of patient mortality (HR 0.92, 95% CI 0.90-0.94, p < 0.001) compared with White recipients.

Among donor factors, a higher KDPI was consistently associated with 68 % higher risk of graft failure (HR 1.68, 95% CI 1.57–1.81) and 37% higher risk of mortality (HR 1.37, 95% CI 1.26–1.48; both p < 0.001). Older donor age demonstrated a small but significant association with graft failure (HR 1.00 per year, 95% CI 1.00–1.00, p < 0.001) and patient mortality (HR 1.00 per year, 95% CI 1.00–1.00, p < 0.001). Higher donor serum creatinine was weakly but significantly associated with greater risk of graft failure (HR 0.99 per mg/dL, 95% CI 0.98–1.00, p = 0.027) and neutral association with mortality (HR 1.00, 95% CI 0.99–1.01, p = 0.420). Donor diabetes conferred a higher risk of both graft failure and patient mortality (graft HR 1.05, p = 0.002; patient HR 1.05, p = 0.013). Donors identified as Black in registry data were associated with modestly higher hazards for both outcomes (graft failure HR 1.08, 95% CI 1.05–1.11; mortality HR 1.05, 95% CI 1.02–1.09), whereas Hispanic donor ethnicity was associated with a slightly lower hazard of graft failure (HR 0.96, 95% CI 0.94–0.99, p = 0.006).

Among transplant-related factors, prolonged cold ischemia time showed a small but statistically meaningful increase in risk for both graft failure and patient death (HR ≈ 1.00 per hour, p < 0.001), reflecting cumulative ischemic injury over time. Interestingly, after adjustment, DCD status was associated with slightly lower hazards for graft loss (HR 0.91, 95% CI 0.89–0.93, p < 0.001) and neutral association with patient mortality (HR 1.01, 95% CI 0.98–1.04, p = 0.488). This reversal from the unadjusted Kaplan–Meier results reflects adequate model control for confounding donor and recipient variables. As shown in ***Fig 11*,** the pattern of risk differs between graft failure and patient mortality, with several predictors demonstrating distinct effect sizes for the two endpoints.

**Fig 11.**
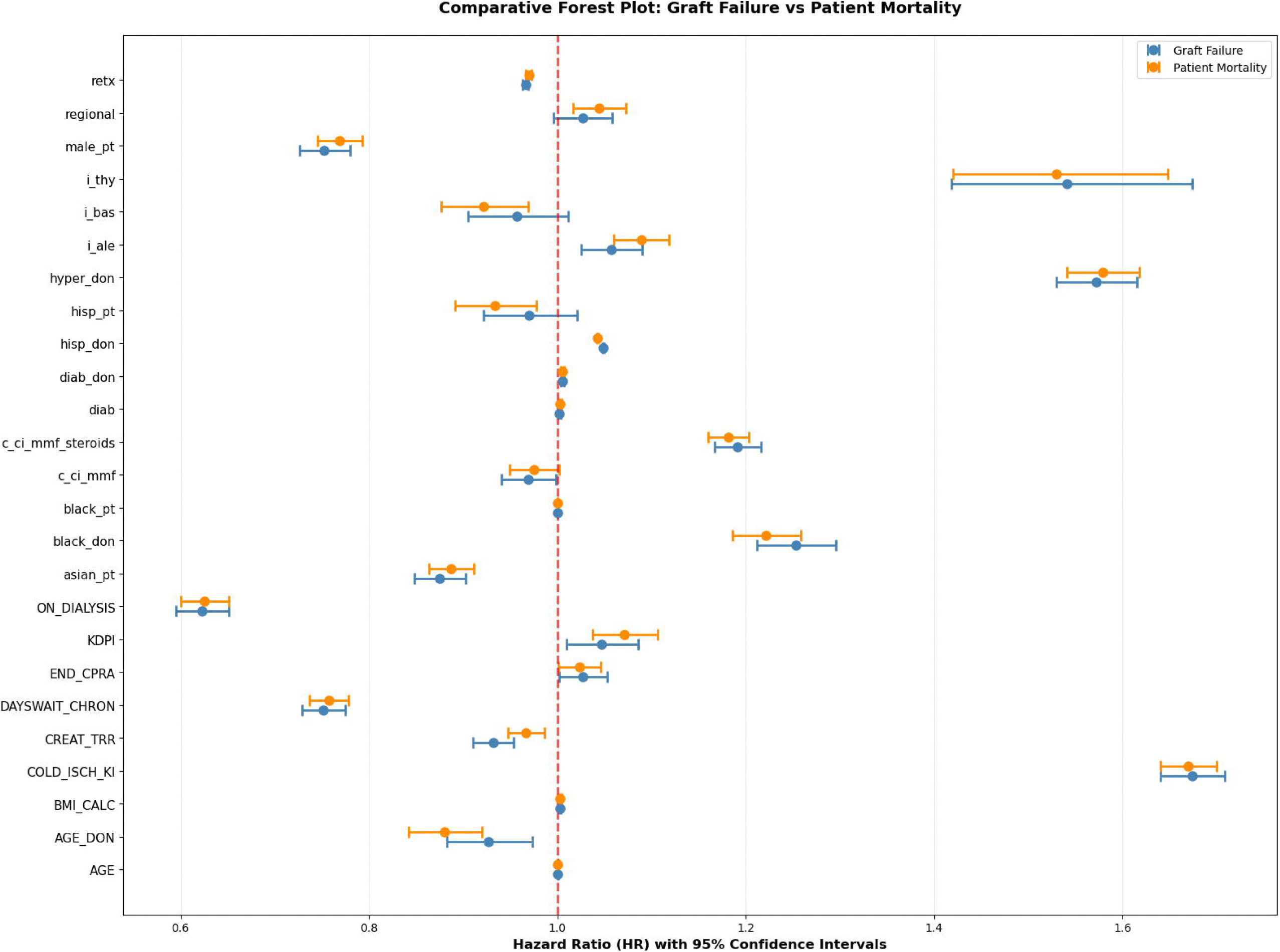
Comparative Forest plot: Graft Failure Vs Patient Mortality. Forest plot depicting adjusted hazard ratios (HRs) and 95% confidence intervals (CIs) for major predictors of graft failure and patient mortality. Values to the left of the vertical reference line (HR = 1) indicate lower risk, and those to the right indicate higher risk. Model adjusted for donor, recipient, and transplant factors.

### Prediction performance

The predictive performance of the classical Cox PH model was benchmarked against four ML survival algorithms using an independent test cohort (***Table 6***). For graft failure, the classical Cox PH model achieved a C-index of 0.687 and mean tdAUC of 0.706, with horizon-specific AUCs of 0.692, 0.688, 0.699, and 0.746 at 1, 3, 5, and 10 years, respectively.

**Table 6.** Model performance comparison for graft failure and patient mortality.

| Model | C-index | tdAUC @ 1 yr | tdAUC @ 3 yr | tdAUC @ 5 yr | tdAUC @ 10 yr | Mean tdAUC |
| --- | --- | --- | --- | --- | --- | --- |
| **Graft Failure** |  |  |  |  |  |  |
| XGBoost-Cox | 0.689 | 0.695 | 0.691 | 0.703 | 0.749 | **0.710** |
| CoxNet | 0.686 | 0.691 | 0.688 | 0.699 | 0.749 | 0.707 |
| Cox PH (Classical) | 0.687 | 0.692 | 0.688 | 0.699 | 0.746 | 0.706 |
| SVM | 0.687 | 0.696 | 0.690 | 0.698 | 0.736 | 0.705 |
| RSF | 0.680 | 0.685 | 0.682 | 0.695 | 0.750 | 0.703 |
| **Patient Mortality** |  |  |  |  |  |  |
| XGBoost-Cox | 0.705 | 0.708 | 0.708 | 0.721 | 0.771 | **0.727** |
| Cox PH (Classical) | 0.703 | 0.707 | 0.706 | 0.719 | 0.770 | 0.726 |
| CoxNet | 0.703 | 0.706 | 0.705 | 0.719 | 0.769 | 0.725 |
| SVM | 0.703 | 0.709 | 0.707 | 0.718 | 0.763 | 0.724 |
| RSF | 0.700 | 0.703 | 0.702 | 0.717 | 0.768 | 0.723 |

Abbreviations: C-index = concordance index; tdAUC = time-dependent area under the curve; Cox PH = Cox proportional hazards model; RSF = Random Survival Forest; SVM = Support Vector Machine; XGBoost-Cox = Extreme Gradient Boosting optimized with the Cox partial likelihood.; CoxNet = L1-penalized Cox regression.

Among the ML survival models, XGBoost-Cox demonstrated the best overall discrimination (C-index = 0.689, mean tdAUC = 0.710), followed closely by CoxNet (C-index = 0.686, mean tdAUC = 0.707) and SVM (C-index = 0.687, mean tdAUC = 0.705). The RSF yielded slightly lower but comparable performance (C-index = 0.680, mean tdAUC = 0.703).

For patient mortality, all models achieved higher concordance than in graft prediction, with C-index values above 0.70. XGBoost-Cox again outperformed other models (C-index: 0.705, mean tdAUC: 0.727), followed closely by the Cox PH (C-index: 0.703, mean tdAUC: 0.726) and CoxNet (C-index: 0.703, mean tdAUC: 0.725). The SVM (C-index: 0.703, mean tdAUC: 0.724) and RSF (C-index: 0.700, mean tdAUC: 0.723) models also showed stable predictive accuracy across time horizons.

## Discussion

In this large, contemporary national cohort of deceased-donor kidney transplant recipients, immunosuppressive regimen selection emerged as an important factor associated with long-term graft and patient survival. Standard maintenance therapy comprising a CNI and MMF, with or without steroids, was consistently associated with lower risks of graft failure and patient mortality, consistent with their widespread use as predominant maintenance strategies in contemporary practice. The CNI + MMF maintenance regimen was associated with lower hazards of both graft failure (HR 0.72) and patient mortality (HR 0.78), supporting prior evidence that CNI and MMF combinations remain central components of contemporary maintenance immunosuppression while balancing rejection prevention with manageable toxicity profiles [27,28]. Similarly, CNI + MMF + steroid triple therapy was associated with lower hazards of graft failure (HR 0.84) and patient mortality (HR 0.90), in line with clinical trials and registry studies that support the use of steroid-containing regimens, particularly in the early post-transplant period for immunologically high-risk recipients [12, 29-31].

Among induction strategies, ATG only induction was associated with consistently lower hazards for both outcomes (HR 0.93), consistent with its common use in high-risk or sensitized recipients where potent T-cell depletion is warranted [10,11,32]. In contrast, IL-2R antagonists showed neutral associations with both graft failure and patient mortality, reflecting their appropriate application in lower-risk populations where the safety profile may outweigh efficacy considerations [8,33,34]. Similarly, Alemtuzumab demonstrated neutral effects, consistent with prior reports indicating that its benefits may be limited to specific induction scenarios [8]. Notably, combination therapy with ATG + IL-2R antagonists was associated with increased hazard for graft loss but not for patient death, which may reflect treatment selection for recipients with greater immunologic complexity or higher baseline risk rather than a direct pharmacologic effect of dual antibody exposure. Given that induction and maintenance regimens reflect underlying immunologic risk and center-specific decision-making rather than random allocation, these findings should be interpreted as adjusted associations rather than causal treatment effects.

Recipient characteristics also exerted substantial influence on long-term outcomes. Diabetes mellitus was the most potent adverse predictor, increasing the hazards of graft loss and mortality by more than 60%. This finding aligns with prior studies linking diabetic nephropathy and systemic metabolic inflammation to accelerated graft loss [18,35,36]. Older age, dialysis dependence, and retransplantation were similarly associated with increased hazards, reflecting cumulative vascular and immunologic injury in these populations [37,38]. In contrast, recipients identified as Asian or Hispanic ethnicity in registry data demonstrated lower adjusted hazards for both graft and patient outcomes relative to White recipients. This observation, replicated in prior registry analyses, may reflect complex interactions between pharmacogenomics, socioeconomic access, and immunologic adaptation [39]. The finding of modestly reduced mortality among Black recipients requires careful interpretation within the context of documented higher immunologic graft loss in this population, suggesting influences from strong selection factors and competing risks.

Among donor characteristics, a higher KDPI was associated with substantially elevated risk of graft failure and mortality, reaffirming the prognostic utility of this composite measure in organ allocation and risk counseling [40,41]. Donor age showed a statistically significant but clinically small association with both graft failure and mortality (HR ≈ 1.00 per year), consistent with donor age being a primary, though not exclusive, component of the KDPI score [40,42]. Donor terminal serum creatinine demonstrated a statistically borderline, inverse association with graft failure (HR 0.99, 95% CI 0.98–1.00, p = 0.027) and a neutral association with patient mortality (HR 1.00, p = 0.420). Although the graft-related association reached statistical significance, the extremely small effect size suggests this finding more likely reflects clinical allocation patterns, such as younger DCD donors with transient creatinine elevation rather than a direct physiologic effect.

Donor diabetes was associated with modestly increased hazards for both outcomes (HR 1.05 each), a finding that is consistent with the established microvascular pathology observed in diabetic donor kidneys [43,44]. The modest effect size likely reflects the heterogeneity of diabetic donor organs; while some studies show excellent outcomes when diabetic donor kidneys are carefully selected [43], others document poorer survival when diabetes is long-standing or accompanied by marked histopathologic injury [44]. The present findings therefore reflect this heterogeneity, indicating a small population-level risk rather than a uniformly detrimental effect. Donors identified as Black in registry data were also associated with slightly higher hazards of graft loss and patient mortality (graft HR 1.08; mortality HR 1.05). In contrast, Hispanic donor ethnicity was associated with a slightly lower hazard of graft failure (HR 0.96, 95% CI 0.94–0.99, p = 0.006). These associations involving donor race should be interpreted within the broader structural and allocation context captured by registry data, as observed differences may reflect organ allocation practices, donor comorbidity patterns, and unmeasured social determinants rather than intrinsic biologic effects.

Transplant-specific factors, particularly prolonged cold ischemia time, were also associated with marginally increased hazards for both endpoints (HR ≈ 1.00 per hour, p < 0.001), underscoring the ongoing relevance of logistical efficiency and rapid organ revascularization in optimizing graft preservation [45,46]. Notably, after adjustment, kidneys from DCD demonstrated slightly lower hazards for graft loss (HR 0.91, 95% CI 0.89–0.93) and a neutral association with patient mortality (HR 1.01, 95% CI 0.98–1.04). This contrasts with unadjusted Kaplan–Meier findings and highlights the role of confounding factors, particularly donor age, ischemia times, and recipient selection in shaping observed outcomes.

Beyond these clinical associations, the present analysis introduces a novel dual-framework modeling approach that integrates classical Cox PH modeling with a suite of ML survival models. Prior transplant survival studies have typically employed either traditional Cox PH regression [17,19] or, more recently, small-scale ML models limited to single outcomes or older datasets [23,25,26,47]. To our knowledge, few national analyses have concurrently applied multiple ML survival algorithms including XGBoost-Cox, CoxNet, SVM, and RSF alongside Cox model in a dataset exceeding 228,000 recipients and extending to 2024. This dual analytical framework provides both interpretability and predictive benchmarking, addressing the interpretability accuracy tradeoff that often limits ML adoption in clinical transplant research [20–22].

Across both outcomes, ML survival models achieved performance metrics comparable to or slightly exceeding the classical Cox model (C-index ≈ 0.69–0.71; mean tdAUC ≈ 0.71–0.73). While the marginal improvement in discrimination was modest, these results represent an improvement in predictive accuracy over earlier national registry analyses, where traditional Cox and ensemble ML approaches typically achieved C-indices of 0.63–0.68 [48,49]. Recent deep learning applications to Scientific Registry of Transplant Recipients (SRTR) data have similarly shown only modest improvements compared to their own baseline Cox models, with reported C-indices of 0.65–0.66 [50]. International efforts such as the Australian Registry study by [51] reported comparable performance for Cox and RSF models (C-index ≈0.67), reinforcing the observation that ML survival algorithms frequently offer incremental rather than transformative gains when applied to clinical registry data. The enhanced model performance observed in the present analysis may reflect both methodological refinement and the use of a dataset extending through 2024, which captures contemporary shifts in donor utilization, immunosuppressive practice, and organ allocation. Collectively, the results affirm the continued value of Cox regression for clinical inference while demonstrating the feasibility and robustness of ML survival models as complementary tools for risk stratification in modern transplant research.

## Strengths and limitations

This study offers several important strengths. It leverages a large, contemporary national cohort spanning 2000–2024, providing exceptional statistical power and broad generalizability across diverse transplant populations. The dual outcome analysis examining both death-censored graft failure and patient mortality provides comprehensive insights into transplant success, allowing a more complete characterization of long-term transplant outcomes than studies limited to a single metric. A major methodological strength is the use of a dual analytical framework that integrates classical Cox PH modeling with multiple ML survival algorithms. This approach enables a direct comparison of inference-driven and data-driven methods, addressing a frequent gap in transplant analytics where interpretability and predictive performance are rarely examined in parallel.

These findings should, however, be interpreted in the context of certain limitations. As a retrospective analysis of national registry data, this study is inherently subject to confounding by indication, as induction and maintenance immunosuppressive regimens are not randomly assigned but instead reflect clinical judgment, immunologic risk, and center-specific practice patterns. Immunosuppressive protocols may vary across transplant centers, and detailed center-level treatment information was not available in the analyzed registry fields. Although comprehensive multivariable adjustment was performed, residual confounding may persist due to unmeasured variables including medication adherence, immunosuppressive drug levels, detailed biopsy findings, and granular center-level practices. Detailed immunologic compatibility metrics such as crossmatch results, desensitization protocols, and ABO compatibility status were not included in the analytic model, and therefore subgroup analyses based on transplant incompatibility could not be performed. Confounding may also arise from treatment selection, particularly for agents preferentially administered to higher-immunologic-risk recipients, such as T-cell–depleting induction or steroid-sparing protocols. Consequently, the observed associations should be interpreted as hypothesis-generating rather than causal effects. Future analyses incorporating propensity score adjustment or causal inference frameworks may further clarify treatment-specific associations.

Furthermore, immunosuppressive therapy was assessed based on the regimen recorded at the time of transplantation, which may not reflect subsequent dose adjustments, medication switches, or discontinuations over the follow-up period. Finally, while the study captures long-term graft and patient survival with high completeness, it does not include other clinically relevant outcomes such as rejection episodes, patient-reported quality of life or specific adverse events related to immunosuppression.

## Conclusion

In this large national analysis of deceased-donor kidney transplant recipients, maintenance regimens incorporating a CNI and MMF either as dual therapy or as part of a steroid-containing triple regimen, were consistently associated with lower hazards of graft failure and patient mortality compared with alternative combinations, consistent with their widespread use as the foundation of modern immunosuppressive therapy. T-cell–depleting induction with ATG was associated with lower hazards for both endpoints, whereas IL-2R antagonists and Alemtuzumab showed neutral associations. These results highlight the dominant influence of maintenance immunosuppression on long-term graft survival and emphasize the need for personalized induction strategies.

Beyond identifying clinical factors associated with long-term outcomes, this study makes several methodological contributions. The comparative modeling framework revealed that the traditional Cox PH model achieved discrimination comparable to ML survival models, while providing superior clinical interpretability through HR estimation. This supports the continued primacy of Cox models for clinical inference in transplantation research, with machine learning serving complementary roles for specific prediction tasks requiring complex feature interactions.

This study advances kidney transplantation analytics through three principal contributions. First, it provides one of the most extensive and temporally comprehensive national evaluations of immunosuppressive therapy to date. Second, it introduces a comparative modeling paradigm that bridges interpretable hazard modeling with predictive ML approaches, offering a practical template for future transplant analytics. Third, it highlights the translational potential of ML survival frameworks to support individualized risk stratification, particularly as transplant registries evolve to include richer clinical and biomarker data.

Future research should build upon these foundations by incorporating time-varying immunosuppressive exposures, therapeutic drug monitoring, center effects, and dynamic clinical variables. Integration of molecular, genomic, and digital biomarkers may ultimately enable the development of precision immunosuppressive strategies and refined prognostic tools that support personalized care throughout the post-transplant lifespan.

## Acknowledgments

The authors acknowledge that this work originated from the doctoral research of Kunle Apanisile under the supervision of Dr. Naoru Koizumi at George Mason University. The study was independently developed and refined by the authors for publication.

## Author contributions

Conceptualization: Kunle Timothy Apanisile, Naoru Koizumi Methodology: Kunle Timothy Apanisile, Hadi El-Amine, Meng-Hao Li, Naoru Koizumi Formal analysis: Kunle Timothy Apanisile, Meng-Hao Li Writing – original draft: Kunle Timothy Apanisile Writing – review & editing: Naoru Koizumi, Hadi El-Amine Supervision: Naoru Koizumi, Hadi El-Amine, Meng-Hao Li

## Ethics Statement

This study used de-identified registry data from the United Network for Organ Sharing (UNOS). Institutional review board approval and informed consent were not required for this secondary analysis of de-identified data.

## Data Availability

The data underlying this study are third-party data obtained from the Organ Procurement and Transplantation Network (OPTN), which is operated by the United Network for Organ Sharing (UNOS). These registry data contain sensitive, de-identified patient-level information and are subject to legal and contractual restrictions that prevent the authors from sharing them publicly. Other qualified researchers may request access to the same dataset directly from UNOS via the OPTN/UNOS data request process (https://unos.org/data/). Access is granted at the discretion of UNOS following submission and approval of a formal data request and execution of a data use agreement; the authors did not have any special access privileges that others would not have. Analytical code used for data preprocessing, statistical analysis, survival modeling, machine learning implementation, and generation of tables and figures is publicly available at: https://github.com/Olukhunlay-hub/kidney-transplant-immunosuppression-survival

## Supporting information

S1 Table. Sensitivity analysis with vs without transplant era adjustment: death-censored graft failure.

S2 Table. Sensitivity analysis with vs without transplant era adjustment: all-cause patient mortality.

S3 Table. Distribution of Kidney Donor Profile Index (KDPI) by Donation After Circulatory Death (DCD) status.

S4 Checklist. STROBE checklist for cohort studies.

