## Supplemental Table 2 for "Immunosuppressive regimens and long-term kidney transplant outcomes: a dual survival modeling framework"

**S2 Table. Sensitivity analysis with vs without transplant era adjustment: all-cause patient mortality.**

| **Covariate** | **Without era adjustment** | | **With era adjustment** | |
| --- | --- | --- | --- | --- |
|  | **HR (95% CI)** | **P-value** | **HR (95% CI)** | **P-value** |
| ATG induction | 0.93 (0.89–0.98) | 0.004 | 0.97 (0.92–1.02) | 0.253 |
| Alemtuzumab induction | 0.96 (0.90–1.01) | 0.103 | 1.01 (0.96–1.07) | 0.685 |
| IL-2R induction | 0.95 (0.91–1.00) | 0.070 | 0.99 (0.94–1.04) | 0.630 |
| IL-2R + ATG induction | 1.02 (0.96–1.09) | 0.470 | 1.08 (1.01–1.15) | 0.026 |
| CNI + MMF (no steroids) | 0.78 (0.76–0.81) | <0.001 | 0.80 (0.78–0.83) | <0.001 |
| CNI + MMF + steroids | 0.90 (0.88–0.93) | <0.001 | 0.92 (0.89–0.95) | <0.001 |
| KDPI (per 10 units) | 1.37 (1.26–1.48) | <0.001 | 1.43 (1.32–1.55) | <0.001 |
| Recipient Age (per year) | 1.05 (1.05–1.05) | <0.001 | 1.05 (1.05–1.05) | <0.001 |
| Male recipient | 1.19 (1.17–1.22) | <0.001 | 1.20 (1.18–1.23) | <0.001 |
| Black recipient | 0.92 (0.90–0.94) | <0.001 | 0.92 (0.90–0.94) | <0.001 |
| Hispanic recipient | 0.76 (0.74–0.78) | <0.001 | 0.76 (0.73–0.78) | <0.001 |
| Retransplant | 1.22 (1.18–1.26) | <0.001 | 1.22 (1.18–1.26) | <0.001 |
| On dialysis at transplant | 1.54 (1.50–1.58) | <0.001 | 1.52 (1.48–1.56) | <0.001 |
| Recipient diabetes | 1.67 (1.63–1.70) | <0.001 | 1.67 (1.63–1.70) | <0.001 |
| DCD donor | 1.01 (0.98–1.04) | 0.488 | 0.99 (0.96–1.02) | 0.558 |

Sensitivity analysis comparing multivariable Cox models with and without adjustment for transplant era. Transplant era categories were 2000–2004 (reference), 2005–2009, 2010–2014, 2015–2019, and 2020–2024. HRs are from fully adjusted models. Abbreviations: HR, hazard ratio; CI, confidence interval; ATG, antithymocyte globulin; IL-2R, interleukin-2 receptor antagonist; CNI, calcineurin inhibitor; MMF, mycophenolate mofetil; KDPI, Kidney Donor Profile Index; DCD, donation after circulatory death.
