## Supplemental Table 3 for "Immunosuppressive regimens and long-term kidney transplant outcomes: a dual survival modeling framework"

**S3 Table. Distribution of Kidney Donor Profile Index (KDPI) by Donation After Circulatory Death (DCD) status.**

| **Donor Type** | **N** | **Mean KDPI (SD)** | **Median KDPI (IQR)** | **Range** |
| --- | --- | --- | --- | --- |
| DCD | 50,907 | 50.5% (22.8) | 50.0% (33.0–68.0) | 1.0–100.0% |
| Non-DCD | 177,948 | 39.6% (26.2) | 37.0% (17.0–60.0) | 0.0–100.0% |

Values are presented as mean (SD) or median (IQR), as appropriate. KDPI values were obtained from the Scientific Registry of Transplant Recipients and expressed as percentages (0–100%). Group differences were evaluated using the Mann–Whitney U test due to non-normal distribution of KDPI. Abbreviations: KDPI, Kidney Donor Profile Index; DCD, donation after circulatory death; SD, standard deviation; IQR, interquartile range.
